# Primary Resection with Bladder Preservation for Colovesical Fistula: Clinical Outcomes and the Prognostic Significance of Perineural Invasion

**DOI:** 10.64898/2026.05.04.26352423

**Authors:** Peirui Wu, Jianming Yang, Zhenyu Xian, Wenwen Zhong, Li Lu

## Abstract

**Background:** This study evaluated the safety and efficacy of primary resection and anastomosis (PRA) for colovesical fistula (CVF) of diverse etiologies and identified independent prognostic factors for oncological outcomes.

**Methods:** We retrospectively analyzed 112 CVF patients (2017–2024) undergoing PRA with or without a defunctioning stoma, comparing clinical outcomes across benign and malignant cohorts.

**Results:** Benign etiologies accounted for 33.0% (n=37) (colonic diverticulitis (n=19, 51.4%), Crohn’s disease (n=14, 37.8%), and iatrogenic injury (n=4, 10.8%)), all underwent PRA with partial cystectomy, achieving zero mortality and no recurrence. Malignancies (67.0%) primarily included colorectal adenocarcinoma (sigmoid colon cancer (n=44, 58.7%) or rectal cancer (n=31, 41.3%)). Within the malignant cohort, radical cystectomy (n=15) was strictly necessitated by advanced disease features, including distal tumor location and extensive bladder wall invasion (80.0% vs 36.7%, P=0.003). Consequently, this advanced cohort experienced longer operative times (589 vs. 289 min), higher blood loss (600 vs. 100 mL), increased morbidity (80.0% vs. 20.0%, P<0.001), and shorter disease-free survival (DFS) (8 vs. 20 months, P=0.008) compared to those amenable to partial cystectomy (n=60). Crucially, multivariate analysis identified perineural invasion (PNI) (HR: 3.83, 95% CI: 1.49–9.84; P=0.005) as a critical independent predictor of recurrence, reflecting the impact of tumor biology over surgical extent.

**Conclusions:** PRA is a definitive and versatile strategy for CVF. In malignant cases, bladder-preserving strategies are oncologically viable when R0 margins are achievable. Integration of PNI status and neoadjuvant therapy was essential for refining personalized multidisciplinary management.

## 1. Introduction

Colovesical fistula (CVF) is an abnormal epithelialized connection between the colorectum and the bladder. This condition leads to considerable morbidity, primarily manifesting with classic symptoms such as pneumaturia, fecaluria, and recurrent urinary tract infections[1, 2]. Predominantly arising from diverticular disease or advanced colorectal malignancies [3–5] CVF severely compromises patient quality of life and necessitates timely, definitive surgical intervention [6–9].

Historically, the surgical management of CVF has been a subject of contention, particularly regarding the choice between primary resection and anastomosis (PRA) and traditional multi-staged procedures [10, 11]. Recently, the clinical paradigm has shifted toward PRA as the definitive standard of care, often supported by selective defunctioning stomas to safeguard the anastomosis while addressing the fistula in a single operative session[12–14].

In malignant CVF, there are no standardized guidelines, but achieving an R0 resection is paramount. Yet, the optimal extent of bladder involvement, specifically the choice between partial and radical cystectomy, remains poorly defined in terms of functional preservation versus oncological radicality[15, 16]. Furthermore, the prognostic influence of histopathological markers, most notably perineural invasion (PNI)[17, 18], and the downstaging potential of neoadjuvant therapy (NAT) [19, 20] represent critical but underexplored frontiers in pelvic fistula surgery.

To address these evidence gaps, we conducted a comprehensive retrospective analysis of 112 patients undergoing PRA for CVF at a tertiary referral center. This study aims to evaluate the safety of PRA across diverse etiologies and identify the biological determinants that dictate long-term success in malignant CVF.

## 2. Materials and Methods

### 2.1 Study Design and Population

Patients diagnosed with CVF at The Sixth Affiliated Hospital of Sun Yat-sen University were treated by the same group from January 1, 2017, to May 31, 2024, and those presenting with symptoms such as fecaluria, pneumaturia, or recurrent urinary tract infections were enrolled. Detailed medical histories were obtained for all participants.

Inclusion criteria were: 1) radiologically confirmed CVF; 2) availability of complete clinical and pathological records; and 3) adherence to a standardized postoperative follow-up protocol. Exclusion criteria included: 1) loss to follow-up or incomplete data; 2) discrepancy between clinical and final pathological diagnoses; 3) presence of multiple synchronous abdominal malignancies; or 4) comorbid psychiatric disorders or substance abuse precluding reliable participation.

Of 204 screened patients, 112 met the criteria and were included in the final analysis (Figure 1). The cohort was stratified by etiology into a benign group (n=37) and a malignant group (n=75). All patients underwent a PRA procedure, characterized by the definitive excision of the fistulous tract, colorectal resection, and immediate reconstruction (with or without temporary diversion). Within the malignant group, 60 patients underwent partial cystectomy and 15 underwent radical cystectomy.

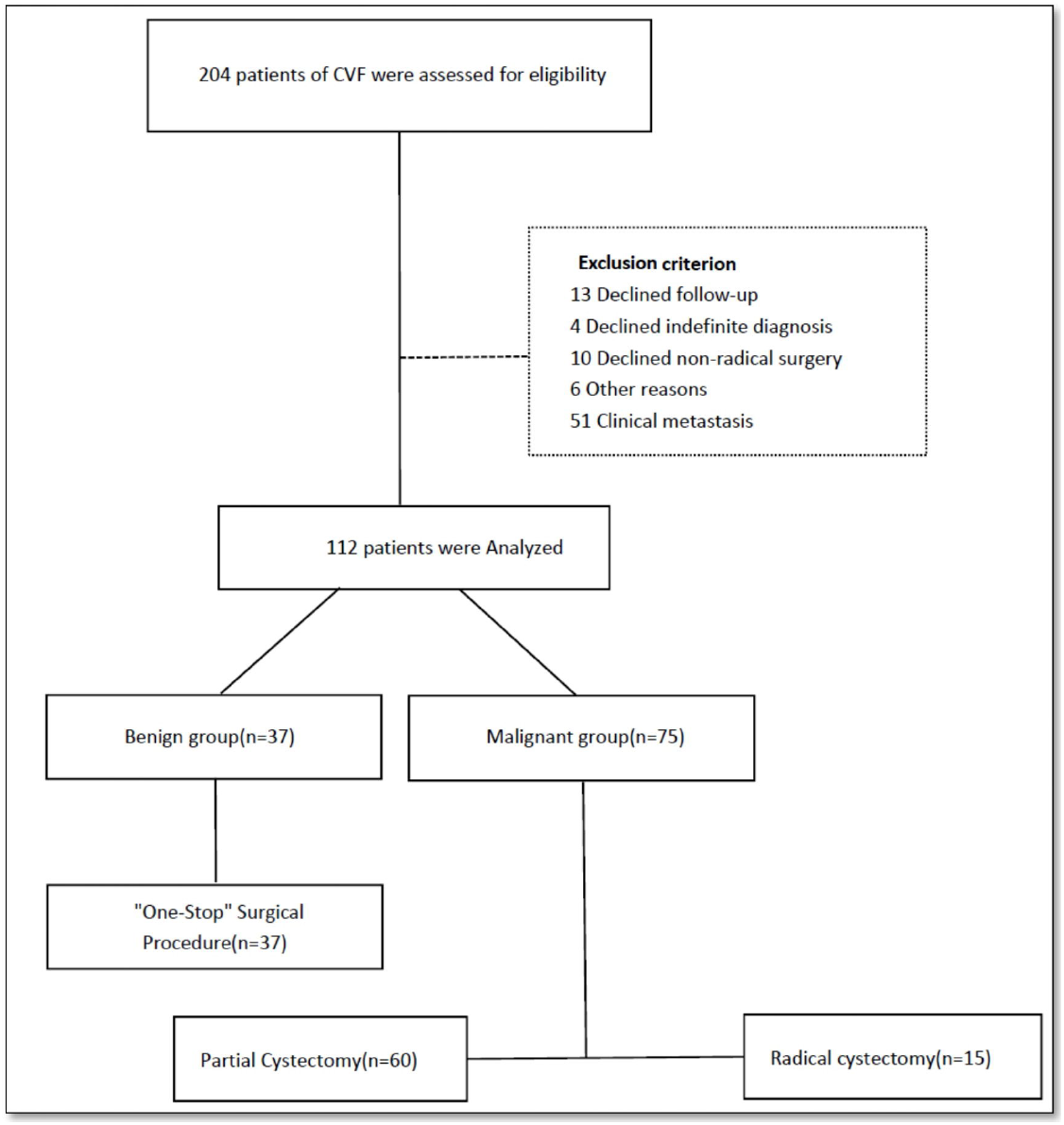

The study was approved by the Ethics Committee of The Sixth Affiliated Hospital of Sun Yat-sen University (2025ZSLYEC-480, 2025-08-29).

### 2.2 Preoperative Assessment and Data Collection

All patients underwent a standardized preoperative evaluation, including routine laboratory tests, contrast-enhanced computed tomography of the chest, abdomen, and pelvis, pelvic magnetic resonance imaging, and colonoscopy and cystoscopy assessments.

Data were systematically extracted from electronic medical records, imaging archives, operative notes, and pathology reports. Collected variables included: 1)Baseline characteristics: age, sex, body mass index, and etiology; 2)Perioperative parameters: surgical approach, operative time, intraoperative blood loss, postoperative complications, length of hospital stay, and reoperation rates; 3)Oncological data (malignant group): Preoperative clinical TNM stage, serum tumor marker levels, and detailed histopathological features from surgical specimens including differentiation grade, lymphovascular invasion, perineural invasion, and circumferential resection margin status. Key radiographic tumor characteristics were also recorded; 4) Surgery safety data: For all patients, complication and mortality data were collected. For malignant cases, oncologic outcomes, including recurrence (local and distant), metastasis, overall survival, and disease-free survival, were documented with precise timelines.

### 2.3 Statistical Analysis

Statistical analyses were performed using SPSS (version 20.0). Continuous data are expressed as mean± standard deviation (if normally distributed) or median with interquartile range (if non-normally distributed). Categorical data are presented as frequencies and percentages. Group comparisons were conducted using appropriate statistical tests: Welch’s t-test or ANOVA for normally distributed continuous variables; the Mann-Whitney U or Kruskal-Wallis tests for non-normally distributed continuous variables; and the Chi-square or Fisher’s exact test for categorical variables.

Survival outcomes were analyzed using the Kaplan-Meier method, and between-group comparisons were assessed using the log-rank test. To identify independent prognostic factors for recurrence and metastasis within the malignant subgroup, variables with a univariate P-value < 0.1 were entered into a multivariate Cox proportional hazards regression model using a stepwise selection procedure. A two-sided P-value < 0.05 was considered statistically significant for all analyses.

## 3. Results

### 3.1 Patient Characteristics and Surgical Management

A total of 112 patients with CVF were included in the analysis, 98 males (87.5%) and 14 females (12.5%), with a median age of 54 years (Table 1). Based on etiology, patients were stratified into a benign group (n=37, 33.0%) and a malignant group (n=75, 67.0%). Benign etiologies included colonic diverticulitis (n=19, 51.4%), Crohn’s disease (n=14, 37.8%), and iatrogenic injury (n=4, 10.8%). In the malignant group, fistulas originated from sigmoid colon cancer (n=44, 58.7%) or rectal cancer (n=31, 41.3%).

All patients underwent PRA, with the specific strategy determined by a multidisciplinary team according to fistula characteristics. For malignant cases, radical colorectal resection with partial cystectomy was performed in 60 patients (80.0%). Radical colorectal resection combined with radical cystectomy was reserved for the remaining 15 patients (20.0%), strictly indicated by advanced disease features such as tumor invasion of the bladder trigone or ureteral orifice, associated hydronephrosis, or a fistula base diameter exceeding 3 cm.

### 3.2 Management and Outcomes of Benign CVF

This subgroup comprised 37 patients, including 34 males (91.9%) and 3 females (8.1%), with a median age of 43 years. 7 patients with relevant comorbidities included peritonitis, and 4 cases with incomplete bowel obstruction.

All procedures were performed under general anesthesia. Patients were placed in the lithotomy position, and cystourethroscopy was first performed to assess the fistula location, its relationship to the ureteral orifices, the degree of local inflammation, and the general condition of the bladder. Abdominal exploration was then undertaken via a laparoscopic five-port approach. The initial focus was the CVF site, followed by a systematic inspection. This included the colon, small bowel, stomach, liver, gallbladder, and spleen. In female patients, the fallopian tubes and uterus were also examined.

PRA achieved 100% success in the benign subgroup, with no recurrences observed during a median 24-month follow-up. Temporary fecal diversion was utilized in 54.1% of cases to protect high-risk anastomoses. The median operative time was 287 minutes with a median blood loss of 100 mL. Postoperative complications occurred in 24.3% of patients, all managed successfully with zero mortality, and the details are as follows:

1. The bladder peritoneum was incised circumferentially, maintaining a minimum margin of 1 cm beyond the CVF, to expose the dome and allow direct excision of the fistulous tract along with the adjacent bladder wall. For smaller fistulas, the bladder and colon were first isolated before proceeding with resection of the fistula and a partial bladder segment.
2. The ureteral orifices were identified and their patency confirmed. Double-J stents were placed when proximity to the resection line posed a risk of injury.
3. The bladder defect was closed in two layers: a full-thickness closure with 2-0 barbed sutures, reinforced by a seromuscular imbricating suture (Supplementary video 1).
4. Following mobilization of the sigmoid mesocolon and upper rectal mesentery with ligation of the inferior mesenteric vessels, the involved colorectal segment was resected. An intracorporeal end-to-end anastomosis was then performed.
5. For high-risk colorectal anastomoses, a diverting ileostomy or loop colostomy was created. A cystostomy was added for cases with precarious bladder repair.

The median operative time was 287 minutes with a median blood loss of 100 mL. One patient (2.7%) required an intraoperative blood transfusion. Temporary fecal diversion (ileostomy or colostomy) was performed in 20 patients (54.1%). Double-J ureteral stents were placed in 20 patients (54.1%), and a cystostomy tube was placed in 11 patients (29.7%).

The median postoperative hospital stay was 10 days. Postoperative complications occurred in 9 patients (24.3%), including Clavien-Dindo grade I-II complications in 4 (10.8%) and grade III-IV in 5 (13.5%). Specific complications were anastomotic leak (n=3, 8.1%), intra-abdominal collection (n=2, 5.4%), parastomal hernia (n=1, 2.7%), bowel obstruction (n=1, 2.7%), and peritonitis (n=1, 2.7%). All complications were managed successfully with no mortality.

During a median follow-up of 24 months, no recurrence of CVF was observed.

### 3.3 Outcomes in Malignant CVF and Comparative Analysis

Following the induction of general anesthesia, patients were placed in the lithotomy position. Cystourethroscopy was first performed to assess the fistula, as previously described. Subsequently, laparoscopic exploration was conducted through five ports. The inspection began at the CVF site, followed by a systematic examination of the colorectum, small bowel, stomach, hepatobiliary system, spleen, and, in female patients, the adnexa and uterus.

All patients underwent a definitive PRA, which consisted of an extended radical colorectal resection combined with either partial or radical cystectomy, as detailed in the following steps:

1. Cystectomy: For partial cystectomy, the peritoneum was incised circumferentially beyond the CVF margin to expose and resect the fistulous tract and the adjacent bladder dome en bloc. For radical cystectomy, the lateral bladder pedicles were mobilized; in male patients, the dorsal venous complex was ligated before division and complete removal of the bladder neck.
2. Urinary Management and Bladder Repair: During partial cystectomy, the patency and proximity of the ureteral orifices were assessed. Double-J stents were placed prophylactically if they were at risk. Bladder closure was performed in two layers: a full-thickness running suture using 2-0 barbed suture, reinforced with a seromuscular imbricating layer.
3. Colorectal Resection and Anastomosis: Following mobilization of the mesocolon and mesorectum, the inferior mesenteric vessels were ligated. The involved colorectal segment was radically excised, and an intracorporeal end-to-end anastomosis was performed (Supplementary video 2).
4. Protective Diversion and Outcomes: A diverting ileostomy or colostomy was created for high-risk colorectal anastomoses; a cystostomy tube was added for tenuous bladder repairs.

#### Disease-Severity Characteristics and Outcomes

The malignant subgroup comprised 75 patients: 60 (80.0%) underwent partial cystectomy and 15 (20.0%) underwent radical cystectomy. No significant differences were observed between the two groups regarding baseline demographics, including age, gender, height, weight, and BMI.

Crucially, the choice of resection extent was governed by disease biology rather than elective preference. Patients requiring radical cystectomy presented with significantly more aggressive local features: lower tumor location (median 10 vs 18 cm, P=0.002) and higher rates of transmural bladder wall invasion (80.0% vs 36.7%, P=0.003).

Consequently, the radical group experienced greater surgical trauma, evidenced by a longer median operative time (589 vs. 289 min, P<0.001) and higher median intraoperative blood loss (600 vs. 100 mL, P<0.001) and significantly shorter disease-free survival (DFS) (8 vs 20 months, P=0.008) (Table 2).

#### Tumor and Pathological Characteristics

There were no significant differences in clinical node staging, tumor thickness, length of bowel involvement, CEA, CA199, CA125, CA153, or the incidence of neoadjuvant therapy between the partial cystectomy and radical cystectomy groups.

Key differences favored the radical cystectomy group for tumors closer to the anal verge (median, 10 vs 18 cm; P=0.002), more frequent rectal origin (60.0% vs 36.7%), and higher pathological rates of bladder wall invasion (80.0% vs 36.7%; P=0.003) and mass-forming morphology (46.7% vs 15.0%; P=0.023). Other pathological features showed no difference.

#### Operative and Postoperative Outcomes

The radical cystectomy group was associated with significantly greater surgical trauma, evidenced by a longer median operative time (589 min [453-715] vs. 289 min [194-358], P<0.001), higher median intraoperative blood loss (600 mL [300-2200] vs. 100 mL [50-200], P<0.001), and a higher intraoperative transfusion rate (53.3% vs. 5.0%, P<0.001). Driven by this advanced disease burden, the radical cystectomy group was associated with significantly greater surgical trauma, evidenced by a longer median operative time (P<0.001), higher median intraoperative blood loss (P<0.001), and a higher overall postoperative complication rate (80.0% vs. 20.0%, P<0.001).

#### Oncologic Outcomes

After a median follow-up of 30 months (IQR, 16-44), reflecting their advanced baseline pathology, the radical cystectomy group had a significantly shorter median disease-free survival (8 months [5–16] vs 20 months [13-42]; P=0.008). No significant intergroup differences were found in median overall survival (19 [7-36] vs 26 [16-44] months; P=0.183), local recurrence rate (26.7% vs 20.0%; P=0.725), or distant metastasis rate (26.7% vs 15.0%; P=0.279).

### 3.4 Survival and Recurrence Analysis among the Malignant Group

Multivariate Cox regression analysis identified specific factors prognostic of oncologic outcomes. For cancer recurrence and metastasis, independent risk factors were rectal location of the CVF (HR: 2.91, 95% CI: 1.10–7.66; P=0.031), necessity of anterior resection plus radical cystectomy (HR: 4.14; P=0.007), and crucially, the presence of PNI (HR: 3.83, 95% CI: 1.49–9.84; P=0.005). PNI was also independently associated with an increased risk of local recurrence (HR: 3.87; P=0.014) (Figure 2B).

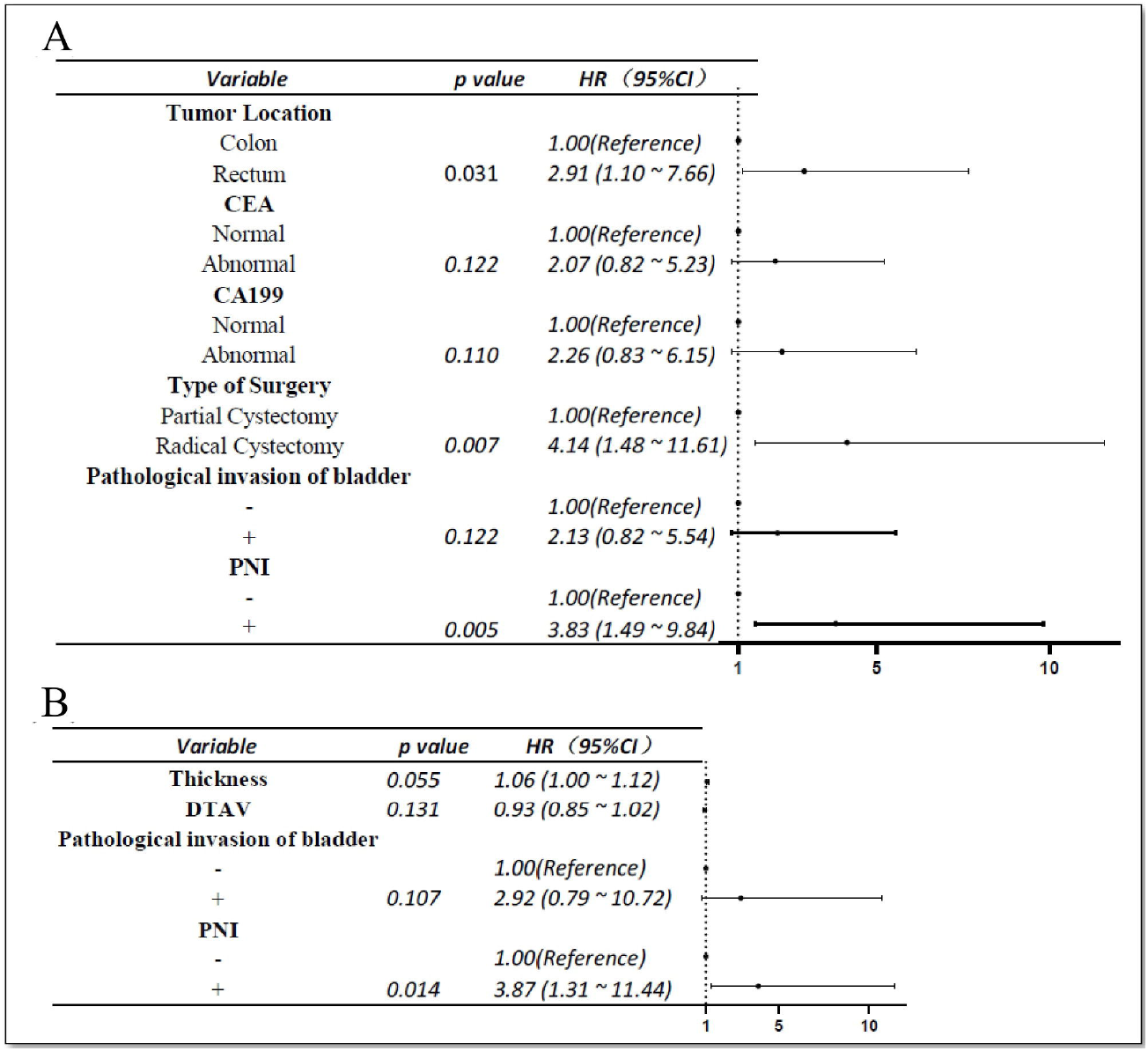

Analysis of overall survival (OS) identified pathological bladder invasion (HR: 8.78, 95% CI: 1.93–39.99; P=0.005) and preoperative metastasis (HR: 6.17, 95% CI: 2.39–15.93; P<0.001) as significant risk factors. In contrast, an ulcerative gross tumor type was a protective factor for OS (HR: 0.04, 95% CI: 0.01–0.22; P<0.001) (Figure 3A). For disease-free survival (DFS), independent predictors of worse outcome were pathological bladder invasion (HR: 3.99, 95% CI: 1.52–10.48; P=0.005), local recurrence (HR: 3.80, 95% CI: 1.70–8.49; P=0.001), and the presence of distant metastasis (HR: 11.31, 95% CI: 4.57–28.00; P<0.001) (Figure 3B).

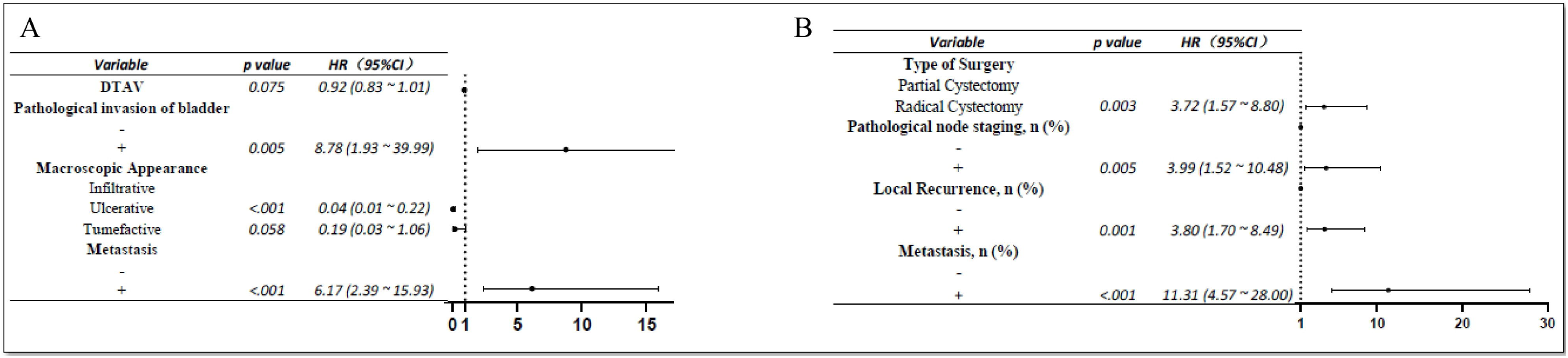

Consistent with these regression findings, Kaplan-Meier analysis confirmed significantly worse survival outcomes in the radical cystectomy group compared to the partial cystectomy group, with shorter overall survival (P=0.0004; Figure 4A) and disease-free survival (P<0.0001; Figure 4B).

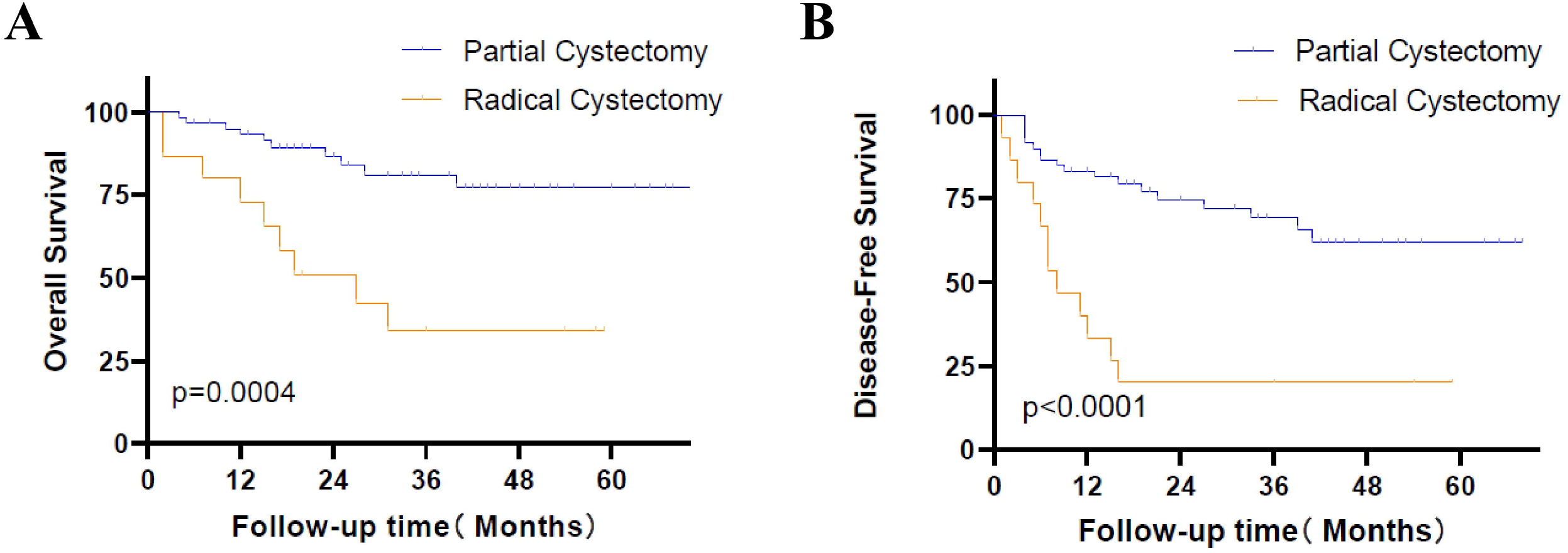

## 4. Discussion

Our findings underscore that a tailored PRA approach is safe and effective across the spectrum of CVF etiologies. For benign diseases, PRA offers a definitive cure with manageable morbidity. In malignant CVF, we emphasize that oncological outcomes are fundamentally determined by tumor biology and anatomical infiltration rather than by surgical choice.

A significant contribution of this study is clarifying the “single-stage” concept. By standardizing on the term PRA, we demonstrate that definitive repair can and should be performed in the primary operation, even when a protective diversion is required. This strategy mitigates the psychological and physical burden associated with multi-stage procedures, such as the classic Hartmann’s operation[10, 12]. In the context of malignant disease, our data initially appeared to suggest that radical cystectomy yields inferior outcomes compared to partial cystectomy. However, this observation must be interpreted through the lens of disease severity rather than surgical efficacy.

Furthermore, our data validate bladder-preserving surgery as a robust oncological strategy when R0 margins are achievable [16, 21]. The inferior outcomes observed in the radical cystectomy group are attributed to advanced disease burden, as evidenced by the high prevalence of distal tumors and extensive bladder invasion in this subset. Still, it remains indispensable for achieving local control in extensive cases[22].

A pivotal finding of this study is the potent prognostic significance of PNI. PNI emerged as a robust independent predictor for both overall recurrence (HR: 3.83) and local recurrence (HR: 3.87) in malignant CVF. While PNI is an established marker of aggressive tumor biology in standard colorectal cancer[17, 18], its specific predictive value in the setting of fistulizing pelvic malignancies has been sparsely documented. The presence of PNI suggests that microscopic tumor dissemination along nerve sheaths extends beyond standard resection planes. Consequently, PNI-positive CVF patients warrant highly aggressive adjuvant systemic therapies and intensified postoperative surveillance protocols to mitigate their distinctively high recurrence risk.

Given that radical cystectomy carries substantial morbidity and functional detriment, the integration of NAT offers a promising avenue for downstaging borderline-resectable tumors involving the bladder trigone[19, 20]. While NAT utilization in our cohort did not show a statistical difference in early outcomes, the conceptual shift toward induction chemoradiotherapy to facilitate bladder-preserving surgery (i.e., converting a mandatory radical cystectomy into a partial cystectomy) represents a crucial evolution in treating locally advanced malignant CVF. Future multidisciplinary algorithms must prioritize identifying NAT-responsive phenotypes.

The interpretation of our findings must consider several limitations inherent to the study’s retrospective, single-center design. Potential selection bias and the influence of institutional expertise may affect the generalizability of our results. Specifically, the inherent disparities in baseline tumor staging between the partial and radical cystectomy cohorts limit direct surgical comparisons, though they accurately reflect real-world clinical triage. Although we reported a median follow-up of 30 months, a longer observation period is required to fully assess long-term oncologic outcomes and quality of life[1, 23].

PRA is a safe, effective, and definitive intervention for CVF. In malignancies, a tailored approach favoring bladder preservation maintains oncological control while reducing morbidity. The integration of PNI as a prognostic biomarker and the strategic use of NAT are essential for optimizing long-term survival and quality of life in this complex patient population.

## Data Availability

The data that support the findings of this study are available from the corresponding author upon reasonable request, subject to institutional ethical approval

## Author Contribution Statement

Peirui Wu: Conceptualization, Formal analysis, Writing-original draft, Writing-review and editing.

Jianming Yang: Conceptualization, Formal analysis, Writing-original draft.

Zhengyu Xian: Conceptualization, Data curation, Investigation.

Wenwen Zhong: Data curation, Investigation, MDT consultation.

Li Lu: Conceptualization, Methodology, Formal analysis, Writing-original draft, Writing-review and editing.

## Notes

### Competing Interest Statement

The authors have declared no competing interest.

### Funding Statement

This study did not receive any funding

## References

1. Kavanagh D, Neary P, Dodd JD, Sheahan KM, O’Donoghue D, Hyland JM: Diagnosis and treatment of enterovesical fistulae. Colorectal Dis 2005, 7(3):286–291.

2. Golabek T, Szymanska A, Szopinski T, Bukowczan J, Furmanek M, Powroznik J, Chlosta P: Enterovesical fistulae: aetiology, imaging, and management. Gastroenterol Res Pract 2013, 2013:617967.

3. Garcea G, Majid I, Sutton CD, Pattenden CJ, Thomas WM: Diagnosis and management of colovesical fistulae; six-year experience of 90 consecutive cases. Colorectal Dis 2006, 8(4):347–352.

4. Hechenbleikner EM, Buckley JC, Wick EC: Acquired rectourethral fistulas in adults: a systematic review of surgical repair techniques and outcomes. Dis Colon Rectum 2013, 56(3):374–383.

5. Melchior S, Cudovic D, Jones J, Thomas C, Gillitzer R, Thuroff J: Diagnosis and surgical management of colovesical fistulas due to sigmoid diverticulitis. J Urol 2009, 182(3):978–982.

6. Diamantidis D, Papatheodorou N, Kostoglou P, Tsakaldimis G, Botaitis S: Management of vesicoenteric fistulas arising from perforated Meckel’s diverticulum: a report of a case and review of the literature. Oxf Med Case Reports 2024, 2024(2):omad155.

7. Moss RL, Ryan JA, Jr.: Management of enterovesical fistulas. Am J Surg 1990, 159(5):514–517.

8. Pontari MA, McMillen MA, Garvey RH, Ballantyne GH: Diagnosis and treatment of enterovesical fistulae. Am Surg 1992, 58(4):258–263.

9. Najjar SF, Jamal MK, Savas JF, Miller TA: The spectrum of colovesical fistula and diagnostic paradigm. Am J Surg 2004, 188(5):617–621.

10. Halim H, Askari A, Nunn R, Hollingshead J: Primary resection anastomosis versus Hartmann’s procedure in Hinchey III and IV diverticulitis. World J Emerg Surg 2019, 14:32.

11. Biffoni M, Urciuoli P, Grimaldi G, Eberspacher C, Santoro A, Pironi D, Sorrenti S: Colovesical fistula complicating diverticular disease: diagnosis and surgical management in elderly. Minerva Chir 2019, 74(2):187–188.

12. Pau S, Patel A, Yap S, Eglinton T, Fischer J: Colovesical Fistula Management and the Role of Cystoscopy: A Single Institution Experience. ANZ J Surg 2025, 95(10):2148–2154.

13. Dong C, Pan X, Wei L, Man X, Zhou Z, Huang Y, Wang X, Qi L, Xue F, Li Y: Colon cancer with colovesical fistula: A report of four cases and a literature review. Oncol Lett 2023, 25(4):158.

14. Zizzo M, Tumiati D, Bassi MC, Zanelli M, Sanguedolce F, Porpiglia F, Fiori C, Campobasso D, Castro Ruiz C, Bergamaschi FA et al: Management of colovesical fistula: a systematic review. Minerva Urol Nephrol 2022, 74(4):400–408.

15. Badic B, Leroux G, Thereaux J, Joumond A, Gancel CH, Bail JP, Meurette G: Colovesical Fistula Complicating Diverticular Disease: A 14-Year Experience. Surg Laparosc Endosc Percutan Tech 2017, 27(2):94–97.

16. Nakamori S, Kawai K, Dejima A, Natsume S, Ise I, Kato H, Takao M, Nakano D: Surgical outcomes of a partial or total cystectomy for colorectal cancer invasion of the bladder. Asian J Surg 2024.

17. Liebig C, Ayala G, Wilks J, Verstovsek G, Liu H, Agarwal N, Berger DH, Albo D: Perineural invasion is an independent predictor of outcome in colorectal cancer. J Clin Oncol 2009, 27(31):5131–5137.

18. Cienfuegos JA, Martinez P, Baixauli J, Beorlegui C, Rosenstone S, Sola JJ, Rodriguez J, Hernandez-Lizoain JL: Perineural Invasion is a Major Prognostic and Predictive Factor of Response to Adjuvant Chemotherapy in Stage I-II Colon Cancer. Ann Surg Oncol 2017, 24(4):1077–1084.

19. Sauer R, Becker H, Hohenberger W, Rodel C, Wittekind C, Fietkau R, Martus P, Tschmelitsch J, Hager E, Hess CF et al: Preoperative versus postoperative chemoradiotherapy for rectal cancer. N Engl J Med 2004, 351(17):1731–1740.

20. Garcia-Aguilar J, Patil S, Gollub MJ, Kim JK, Yuval JB, Thompson HM, Verheij FS, Omer DM, Lee M, Dunne RF et al: Organ Preservation in Patients With Rectal Adenocarcinoma Treated With Total Neoadjuvant Therapy. J Clin Oncol 2022, 40(23):2546–2556.

21. Pokala N, Delaney CP, Brady KM, Senagore AJ: Elective laparoscopic surgery for benign internal enteric fistulas: a review of 43 cases. Surg Endosc 2005, 19(2):222–225.

22. PelvEx C: Factors affecting outcomes following pelvic exenteration for locally recurrent rectal cancer. Br J Surg 2018, 105(6):650–657.

23. Bahadursingh AM, Longo WE: Colovaginal fistulas. Etiology and management. J Reprod Med 2003, 48(7):489–495.

