## Supplementary material for "Primary Resection with Bladder Preservation for Colovesical Fistula: Clinical Outcomes and the Prognostic Significance of Perineural Invasion": Table 1 and 2

Table 1. Clinical characteristics and surgery related parameters (n=112).

| Characteristic | n = 112 |
| --- | --- |
| Age (years), Mean ± SD | 54 ± 15 |
| Gender, n (%) |  |
| Male | 98 (87.5%) |
| Female | 14 (12.5%) |
| Etiology, n (%) |  |
| Iatrogenic injury | 4 (3.5%) |
| Colonic diverticula | 19 (17.0%) |
| Crohn's disease | 14 (12.5%) |
| Colorectal cancer | 75 (67.0%) |
| Surgery time（mins）, Median (Q1, Q3) | 308 (221, 443) |
| Intraoperative blood loss（ml）, Median (Q1, Q3) | 100 (50, 300) |
| Intraoperative blood transfusion, n (%) |  |
| No | 100 (89.3%) |
| Yes | 12 (10.7%) |
| Post-op LOS (days), Median (Q1, Q3) | 10.3 (8.0, 13.9) |
| Clavien-Dindo stage，n (%) |  |
| No | 79 (70.5%) |
| I-II | 14 (12.5%) |
| III-IV | 19 (17.0%) |
| Duration of catheterization (days), Median (Q1, Q3) | 11.0 (8.0, 14.0) |
| Cystostomy, n (%) |  |
| Partial cystectomy | 97 (86.6%) |
| Radical cystectomy | 15 (13.4%) |
| Ostomy, n (%) |  |
| No | 55 (49.1%) |
| Yes | 57 (50.9%) |
| Positions of Ostomy, n (%) |  |
| None | 55 (49.1%) |
| Colon | 24 (21.4%) |
| Ileum | 31 (27.7%) |
| Both | 2 (1.8%) |
| Length of stay (days), Median (Q1, Q3) | 49 (27, 62) |
| Follow-up (months), Median (Q1, Q3) | 27 (14, 44) |

Post-op LOS: Postoperative length of stay.

Table 2. Clinical characteristics and surgery related parameters of malignant CVF(n=75)

| Characteristic | Type of Surgery | | p-value |
| --- | --- | --- | --- |
|  | Partial Cystectomy | Radical Cystectomy |  |
|  | n = 60 | n = 15 |  |
| Age (years), Mean ± SD | 57 ± 15 | 61 ± 10 | 0.212 |
| Gender, n (%) |  |  | 0.683 |
| Male | 52 (86.7%) | 12 (80.0%) |  |
| Female | 8 (13.3%) | 3 (20.0%) |  |
| Height (cm), Mean ± SD | 166 ± 6 | 167 ± 8 | 0.499 |
| Weight (kg), Mean ± SD | 60 ± 8 | 61 ± 9 | 0.710 |
| BMI (kg/m^2^), Mean ± SD | 21.90 ± 2.68 | 21.84 ± 2.43 | 0.929 |
| Clinical node staging, n (%) |  |  | 0.639 |
| 0 | 10 (16.7%) | 3 (20.0%) |  |
| 1 | 25 (41.7%) | 4 (26.7%) |  |
| 2 | 25 (41.7%) | 8 (53.3%) |  |
| Thickness of tumor (mm), Median (Q1, Q3) | 17 (13, 21) | 20 (14, 20) | 0.618 |
| Length of bowel involved (mm), Median (Q1, Q3) | 57 (45, 74) | 49 (40, 64) | 0.162 |
| DTAV (cm), Median (Q1, Q3) | 18 (11, 24) | 10 (4, 15) | 0.002 |
| Tumor Location, n (%) |  |  | 0.101 |
| Colon | 38 (63.3%) | 6 (40.0%) |  |
| Rectum | 22 (36.7%) | 9 (60.0%) |  |
| CEA, n (%) |  |  | 0.419 |
| Normal | 29 (48.3%) | 9 (60.0%) |  |
| Abnormal | 31 (51.7%) | 6 (40.0%) |  |
| CA199, n (%) |  |  | 0.722 |
| Normal | 48 (80.0%) | 13 (86.7%) |  |
| Abnormal | 12 (20.0%) | 2 (13.3%) |  |
| CA125, n (%) |  |  | >0.999^2^ |
| Normal | 57 (95.0%) | 14 (93.3%) |  |
| Abnormal | 3 (5.0%) | 1 (6.7%) |  |
| CA153, n (%) |  |  | >0.999^2^ |
| Normal | 58 (96.7%) | 15 (100.0%) |  |
| Abnormal | 2 (3.3%) | 0 (0.0%) |  |
| Neoadjuvant Therapy, n (%) |  |  | 0.414 |
| No | 27 (45.0%) | 5 (33.3%) |  |
| Yes | 33 (55.0%) | 10 (66.7%) |  |
| Surgery time (mins), Median (Q1, Q3) | 289 (194, 358) | 589 (453, 715) | <0.001^3^ |
| Intraoperative blood loss (ml), Median (Q1, Q3) | 100 (50, 200) | 600 (300, 2,200) | <0.001^3^ |
| Intraoperative blood transfusion, n (%) |  |  | <0.001^2^ |
| No | 57 (95.0%) | 7 (46.7%) |  |
| Yes | 3 (5.0%) | 8 (53.3%) |  |
| Clavien-Dindo stage，n (%) |  |  | <0.001^2^ |
| No | 48 (80.0%) | 3 (20.0%) |  |
| I-II | 5 (8.3%) | 4 (26.7%) |  |
| III-IV | 7 (11.7%) | 8 (53.3%) |  |
| Ostomy, n (%) |  |  | 0.644 |
| No | 32 (53.3%) | 7 (46.7%) |  |
| Yes | 28 (46.7%) | 8 (53.3%) |  |
| Post-op LOS (days), Median (Q1, Q3) | 9.1 (8.0, 12.6) | 14.5 (11.6, 26.6) | <0.001^3^ |
| Pathological invasion of bladder, n (%) |  |  | 0.003 |
| No | 38 (63.3%) | 3 (20.0%) |  |
| Yes | 22 (36.7%) | 12 (80.0%) |  |
| Macroscopic Appearance, n (%) |  |  | 0.023 |
| Infiltrative | 4 (6.7%) | 1 (6.7%) |  |
| Ulcerative | 47 (78.3%) | 7 (46.7%) |  |
| Tumefactive | 9 (15.0%) | 7 (46.7%) |  |
| Pathological node staging, n (%) |  |  | >0.999^2^ |
| No | 44 (73.3%) | 11 (73.3%) |  |
| Yes | 16 (26.7%) | 4 (26.7%) |  |
| Tumor Grade, n (%) |  |  | 0.427 |
| Well-differentiated | 3 (5.0%) | 1 (6.7%) |  |
| Moderately-differentiated | 50 (83.3%) | 11 (73.3%) |  |
| Poorly-differentiated | 7 (11.7%) | 3 (20.0%) |  |
| LVI, n (%) |  |  | 0.408 |
| No | 53 (88.3%) | 12 (80.0%) |  |
| Yes | 7 (11.7%) | 3 (20.0%) |  |
| PNI, n (%) |  |  | >0.999^2^ |
| No | 47 (78.3%) | 12 (80.0%) |  |
| Yes | 13 (21.7%) | 3 (20.0%) |  |
| MMR, n (%) |  |  | 0.578 |
| dMMR | 56 (93.3%) | 15 (100.0%) |  |
| pMMR | 4 (6.7%) | 0 (0.0%) |  |
| Local recurrence, n (%) |  |  | 0.725 |
| No | 48 (80.0%) | 11 (73.3%) |  |
| Yes | 12 (20.0%) | 4 (26.7%) |  |
| Distant metastasis, n (%) |  |  | 0.279 |
| No | 51 (85.0%) | 11 (73.3%) |  |
| Yes | 9 (15.0%) | 4 (26.7%) |  |
| OS, Median (Q1, Q3) | 26 (16, 44) | 19 (7, 36) | 0.183 |
| DFS, Median (Q1, Q3) | 20 (13, 42) | 8 (5, 16) | 0.008 |
| ^1^Welch Two Sample t-test; ^2^Fisher's exact test; ^3^Wilcoxon rank sum test;^4^Pearson's Chi-squared test; | | | |
| DTAV: Distance from the anal verge; MMR: Mismatch repair; LVI: Lymphovascular invasion; PNI: Perineural invasion; Post-op LOS: Postoperative Length of Stay; OS: Overall survival; DFS: Disease-Free Survival. | | | |
